# Genetic liability to ADHD and migraine risk: Mendelian randomization evidence for behavioural and psychiatric pathways

**DOI:** 10.64898/2026.09.08.26362527

**Authors:** Yaxin Luo, Christina Dardani, Robyn E Wootton, Eleanor C M Sanderson, Evie Stergiakouli

## Abstract

**Background:** Attention-deficit/hyperactivity disorder (ADHD) and migraine frequently co-occur, but the mechanisms underlying this association remain unclear. Examining migraine with aura and without aura separately may help clarify whether ADHD-related pathways are shared across migraine subtypes or differ by aura status.

**Methods:** Two-sample Mendelian randomization (MR) was used to examine the effect of genetic liability to ADHD on overall migraine and on migraine subtypes defined by aura status. MR-based mediation was then used to assess whether behavioural, psychiatric, sleep-related, education, and cardiometabolic traits contribute to these effects.

**Results:** MR provided evidence for an effect of ADHD genetic liability on overall migraine risk (OR = 1.48, 95% CI =1.2, 1.82), with similar effect sizes for migraine with aura (MA) (OR=1.52, 95% CI = 1.18, 1.96) and migraine without aura (MO) (OR = 1.65, 95% CI = 1.19, 2.31). Depression, alcohol intake frequency, lifetime smoking index, ever smoke, risk taking behaviours, insomnia, and college education showed evidence for partial mediation, explaining approximately 34.17% (95% CI = 6.17%, 61.63%), 14.44% (95% CI = 0%, 28.87%), 37.25% (95% CI = 11.86%, 63.64%), and 29.24% (95% CI = 6.12%, 52.37%) of the total effect, individually. Subtype analyses suggested broadly similar patterns across MA and MO, with no clear subtype-specific differences.

**Conclusion:** Our findings provide evidence that ADHD common variant genetic liability increases migraine risk, including both MA and MO. The association appears unlikely to be explained by a single pathway and may involve the contribution of depression-related, lifestyle-related, and education-related processes.

## Introduction

Attention-deficit/hyperactivity disorder (ADHD) and migraine are both common, heritable conditions that contribute substantially to global disease burden across the life course (1, 2). ADHD is a neurodevelopmental condition characterized by persistent patterns of inattention and/or hyperactivity-impulsivity. Onset is typically in childhood, with a substantial proportion of individuals experiencing persistence into adulthood (3–5). Migraine is a recurrent primary headache disorder, often accompanied by sensory and autonomic symptoms, affecting up to 15% of adults worldwide and is a leading cause of disability, particularly among women of working age (6). Although ADHD and migraine differ in clinical presentation and typical age at onset, epidemiological studies suggest that they commonly co-occur (7).

Observational, longitudinal and genetic studies have reported evidence linking ADHD and migraine. Cross-sectional studies suggested higher migraine prevalence among individuals with ADHD, compared with those without ADHD (8, 9). Longitudinal evidence from a Taiwanese health insurance cohort found that individuals with ADHD had an increased risk of developing migraine compared with matched controls (Hazard ratio = 1.92, 95% CI 1.64, 2.34) (10). Mendelian randomization (MR) analysis has also provided evidence consistent with a potential effect of ADHD liability on migraine risk, although the estimated effect was modest (OR_IVW_ = 1.08, 95% CI: 1.02, 1.13) (11). However, the pathways underlying this association remain unclear.

Migraine comprises clinically distinct subtypes that may not have identical relationships with ADHD liability. Migraine with aura (MA) is characterised by transient and fully reversible neurological symptoms, most often visual or sensory symptoms, whereas migraine without aura (MO) occurs in the absence of these focal aura symptoms (12). Family studies suggested that familial aggregation may differ by aura status, with MA showing a stronger genetic component than MO (13). Some subtype-specific loci may point to biologically distinct mechanisms (14); for example, the MA-specific signal near *CACNA1A* is consistent with neuronal calcium-channel function and cortical excitability, processes that are relevant to aura (15). However, environmental and lifestyle-related factors, including stress, sleep disruption, hormonal changes, smoking, and alcohol intake are involved in both migraine subtypes.

ADHD is associated with several behavioural, psychiatric, sleep-related, and cardiometabolic traits that may also influence migraine susceptibility, including smoking, alcohol use, sleep disturbance, depression, body mass index (BMI) (16, 17). These traits could lie on downstream pathways from ADHD liability to migraine risk (9, 18). If ADHD genetic liability has an effect on migraine, it might be mediated through downstream behavioural, psychiatric, sleep-related, or cardiometabolic traits. Identifying these pathways may help clarify the mechanisms underlying the association and whether they differ between MA and MO. Nevertheless, establishing such pathways using conventional observational analyses is challenging because associations across the proposed mediation pathway may be influenced by residual confounding or overlapping genetic liability (17, 19).

We examined whether genetic liability to ADHD was associated with overall migraine and migraine subtype defined by aura status and whether the mediating effects of behavioural, psychiatric, sleep-related, and cardiometabolic traits were shared across the two subtypes or differed between them. MR-based mediation analyses can help distinguish the total effect of an exposure from pathways operating through intermediate traits (20). We first used univariable MR to estimate the overall effect of genetic liability to ADHD on overall migraine (21). Two-step MR was then used to assess whether ADHD liability was associated with each candidate mediator and whether genetic liability to the mediator was subsequently associated with migraine. Where both steps were supported, multivariable MR was used to estimate the direct effect of ADHD liability on migraine after accounting for the mediator.

## Methods

Mendelian randomization analyses

### Data sources

#### Exposure: genetic liability to ADHD

Genetic instruments for ADHD were obtained from the most recent GWAS meta-analysis of ADHD, which combined clinically diagnosed ADHD and quantitative ADHD symptom scores across 296,487 participants of European ancestry (22). The GWAS identified 2,039 genome-wide significant SNPs (P<5×10^-8^), corresponding to 43 independent lead variants across 39 loci, and reported a SNP-based heritability of 0.11 (SE = 0.01). Genetic instruments used in the present MR analyses were selected from the GWAS summary statistics according to the criteria described below.

#### Candidate mediators

Candidate mediators were selected based on three criteria informed by previous observational and genetic literature. First, there needed to be evidence that the trait was associated with both ADHD and migraine. Second, the trait needed to be biologically or behaviourally plausible as a downstream pathway from ADHD liability to migraine. Third, suitable GWAS data had to be available for MR analysis.

We therefore focused on behavioural, lifestyle, sleep-related, psychological, and cardiometabolic pathways that may link ADHD liability to migraine risk. Smoking behaviour was represented primarily by lifetime smoking index, which reflects cumulative smoking exposure based on smoking initiation, duration, heaviness, and cessation (23). Smoking initiation was also included as a secondary smoking phenotype because of its widespread use in genetic studies and its comparability with previous MR analyses (24).

Sleep-related traits include insomnia, sleep duration, and overall sleep health score, reflecting evidence linking sleep disturbance to migraine risk, although evidence is more consistent for insomnia than for sleep duration (25, 26). Additional candidate mediators include physical activity, alcohol intake frequency, chronotype, college education, and risk-taking behaviour (27–30).

For some mediators, different GWAS sources were used across analysis steps because data availability differed. Full summary statistics were required for the ADHD-mediator step and MVMR analyses, as these required associations estimates for ADHD instruments or pooled ADHD-mediator instruments. GWAS sources reporting only top associated variants were used only for univariable mediator-to-migraine analyses, where these variants served as mediator instruments. GWAS sources and sample sizes are presented separately for mediators analysed as outcomes in the ADHD-to-mediator step (Table S1) and as exposures in the mediator-to-migraine step (Table S2), with instrument numbers reported for the exposure datasets.

#### Outcome: GWAS of overall migraine and subtype migraine

Migraine GWAS summary statistics were obtained from FinnGen release 13. Migraine was defined using clinical diagnoses recorded in Finnish national health registries. The overall migraine GWAS included 28,504 cases and 366,556 controls. The MA GWAS included 12,743 cases and 366,556 controls, and the MO GWAS included 10,206 cases and 366,556 controls (31). Although larger migraine GWAS datasets are available, several include a substantial UK Biobank contribution and were therefore not used as the outcome dataset to minimise sample overlap with mediator GWASs which were based on UK Biobank participants (14).

#### Genetic instrument selection

SNPs associated with ADHD at genome-wide significance (P < 5×10^-8^) were selected as candidate instruments and clumped to ensure independence using a linkage disequilibrium threshold of r^2^ < 0.001 within a 10,000kb window, based on 1000 Genomes European reference panel, consistent with the European ancestry of the GWAS population.

#### Mendelian randomisation and mediation analyses

MR was used to estimate the effect of genetically predicted ADHD liability on candidate mediators and migraine. These estimates were interpreted as causal effects under the core instrumental variable (IV) assumptions (32, 33). First, IVs must be strongly associated with the exposure of interest. Second, there should be no confounding of the association between the genetic instrument and the outcome. Third, IVs should affect the outcome only through the exposure and not through other biological pathways. Instrument strength was assessed using the F-statistic, with F > 10 used to indicate sufficient strength (34). SNP-exposure and SNP-outcome associations were harmonised so that effect alleles were aligned across datasets. Palindromic variants with ambiguous allele frequencies were removed. For each SNP, a Wald ratio estimate was calculated as the SNP-outcome association divided by SNP-exposure association. Where multiple instruments were available, these SNP-specific estimates were combined using inverse-variance weighted (IVW) MR as the primary estimator (21).

Multivariable MR (MVMR) was used to estimate the effect of each candidate mediator on migraine after accounting for genetic liability to ADHD. In this framework, genetic variants may be associated with one or more included exposures, but valid instruments should not affect the outcome through pathways other than through the exposures included in the model (35).

For each MVMR analysis, we generated a pooled set of SNPs associated with either ADHD or the candidate mediator at the genome-wide significance level. The pooled SNP set was clumped to account for linkage disequilibrium across traits. SNP associations for ADHD, the mediator, and migraine were then extracted and harmonised across datasets. Instrument strength within each multivariable model was assessed using the conditional F-statistic for summary-data MVMR (36).

#### Prioritisation of candidate mediators for mediation analysis

Candidate mediators were prioritised using a stepwise framework (Figure 1). First, univariable MR (UVMR) was used to assess evidence for both components of the mediation pathway: the effect of genetic liability to ADHD on each candidate mediator, and the effect of each candidate mediator on migraine. Candidate mediators showing evidence for both components were taken forward for mediation-MR analyses.

**Figure 1.**
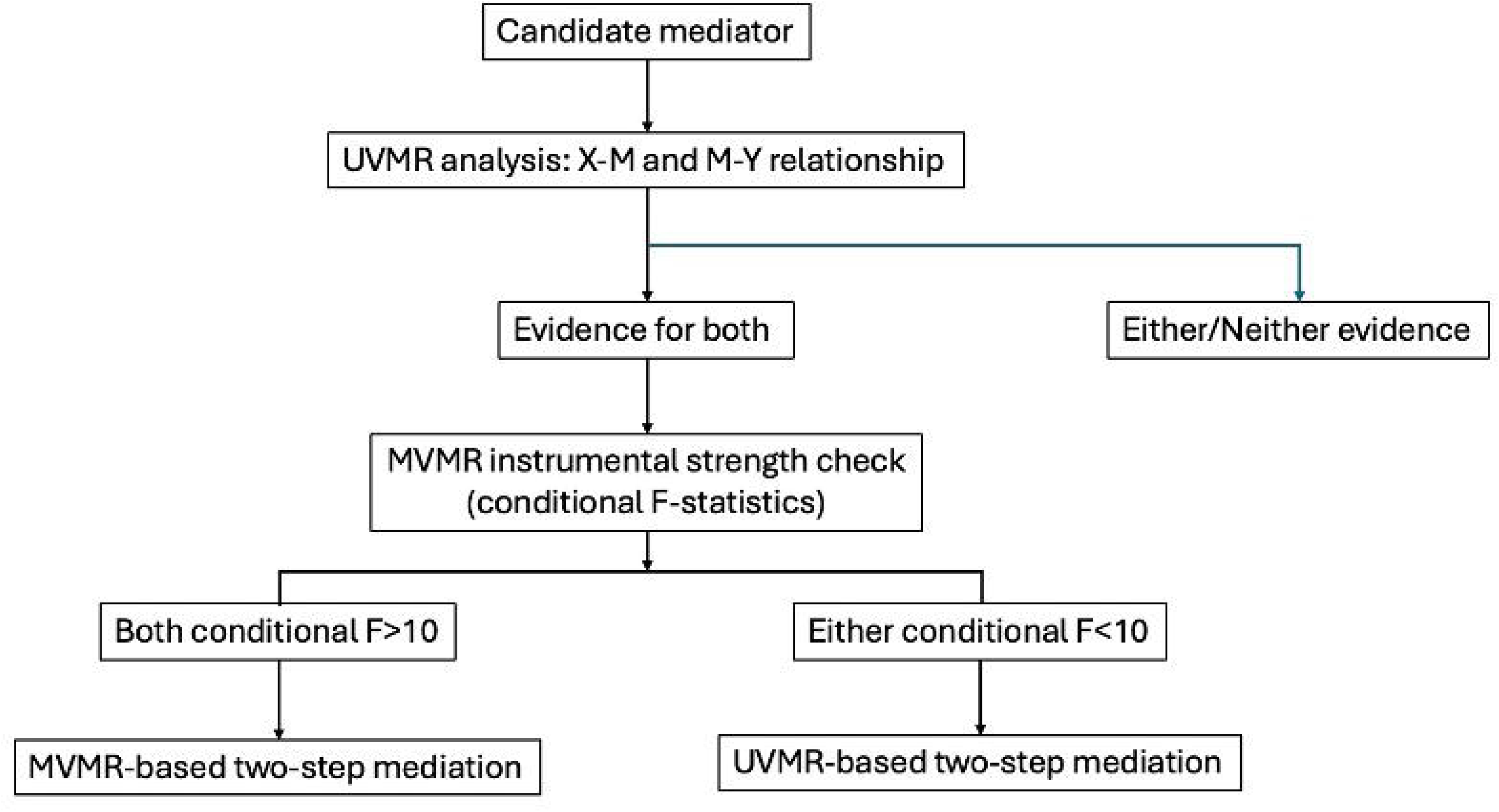
Prioritisation framework for two-step Mendelian randomization mediation analyses based on UVMR evidence and conditional instrument strength.

We then assessed whether each prioritised mediator could be analysed using MVMR. Where full summary statistics were available for the required SNP set, conditional F-statistics were calculated for each ADHD-mediator pair to assess the strength of instruments for ADHD and the mediator within the multivariable model. Where both conditional F-statistics generated were greater than 10, the mediator was considered suitable for MVMR-based mediation analysis (36).Where only top variant associations were available for the mediator, or where either conditional F-statistic was below 10, MVMR was not considered sufficiently instrumented, and UVMR-based two-step mediation was used as complementary evidence (37).

#### MR-based mediation framework

The effect of ADHD genetic liability on migraine through potential mediators was assessed using a two-step MR framework with the product-of-coefficients approach. The total effect (TE) was defined as the effect of genetic liability to ADHD on migraine, estimated using IVW MR as the primary analysis. The ADHD-mediator effect was denoted as *β_XM_*.

For mediators suitable for MVMR, the mediator-migraine effect conditional on genetic liability to ADHD was used as the second step estimate and denoted as *β_MY_*_|*X*_.

The indirect effect (IE) was derived using the product-of-coefficient method: *IE* = *β_XM_* × *β_MY_*_|*X*_. The coefficient for ADHD from the same MVMR model, *β_MY_*_|*X*_, was interpreted as the direct effect of ADHD liability on migraine not mediated through the candidate mediator.

For mediators not suitable for MVMR, the UVMR mediator-migraine effect was used as the second-step estimate, and the UVMR-based two-step mediation was interpreted instead. Standard errors for the indirect effect were calculated using the Delta method. The proportion mediated was defined as IE/TE, with standard errors and 95% confidence intervals also estimated using the Delta method.

#### Sensitivity analysis

Sensitivity analyses were conducted to assess the robustness of the MR estimates to potential violations of the IV assumptions. Weighted median, MR-Egger, and mode-based estimators were used as pleiotropy-robust approaches for UVMR analyses where sufficient instruments were available (38, 39). Heterogeneity across SNP-specific estimates was assessed using Cochran’s Q-statistic (36), and directional horizontal pleiotropy was assessed using the MR-Egger intercept (40). SNP-level MR-Steiger filtering implemented in the ‘TwoSampleMR’ package was used to identify variants that explained more variance in the outcome than in the exposure and were therefore inconsistent with the hypothesised direction of effect (41). MR analyses were repeated after excluding these variants to assess whether results were robust to potential misspecification of the causal direction.

## Results

### The total effect of ADHD genetic liability on migraine

A total of 39 independent SNPs were extracted as the IVs for ADHD (Table S3). We first estimated the causal effect of ADHD genetic liability on migraine using the summary statistics from the latest available ADHD GWAS. Genetic liability to ADHD was associated with migraine (OR_IVW_=1.48, 95% CI=1.20, 1.82) in the MR-IVW analysis. There was evidence of heterogeneity across the genetic instruments (Q-statistic=70). The MR-Egger intercept was close to zero, with little evidence against the null (β =0.01, *P* = 0.19, Table S4). Limited evidence was found for a causal effect of genetic liability to migraine on ADHD (OR_IVW_ = 1.02, 95% CI = 0.98, 1.06, Table S5).

### Causal effects of genetic liability to ADHD on mediators

In the preliminary univariable MR analysis using the IVW method, ADHD genetic liability was associated with an increased risk for depression (β =0.44, 95% CI = 0.30, 0.59; P=3.500×10^-9^) and elevated BMI (β =0.32, 95% CI = 0.18, 0.46; P=1.200×10^-5^). In terms of sleep traits, ADHD genetic liability was associated with increased risk of insomnia (β =0.11, 95% CI = 0.06, 0.17; P=3.490×10^-5^) and was associated with reduced sleep duration (β =-0.10, 95% CI = -0.18, -0.03; P=0.008). In terms of behaviour traits, ADHD genetic liability was linked to heightened risk-taking (β =0.09, 95% CI = 0.06, 0.12; P =3.850×10^-11^), smoking initiation (β =0.11, 95% CI = 0.07, 0.14; P=3.640×10^-9^), lifetime smoking index (β=0.25, 95% CI = 0.20, 0.30; P = 6.838×10^-20^), and alcohol intake frequency (β=0.28, 95% CI = 0.17, 0.39; P= 2.309×10^-5^). Furthermore, ADHD genetic liability was associated with increased physical activity levels (β =0.08, 95% CI = 0.03, 0.13; P=0.003), while demonstrating a strong negative association with obtaining a college education (β =-0.16, 95% CI = -0.21, -0.1; P=4.770×10^-9^). SNP-level MR-Steiger filtering identified and excluded variants that explained more variance in the outcome than in the exposure. MR estimates after Steiger filtering were broadly consistent with the primary analyses (Figure 2). Sensitivity analyses were generally consistent in direction with the primary ADHD-to-mediator estimates, although the precision of estimates varied across methods Table S6.

**Figure 2.**
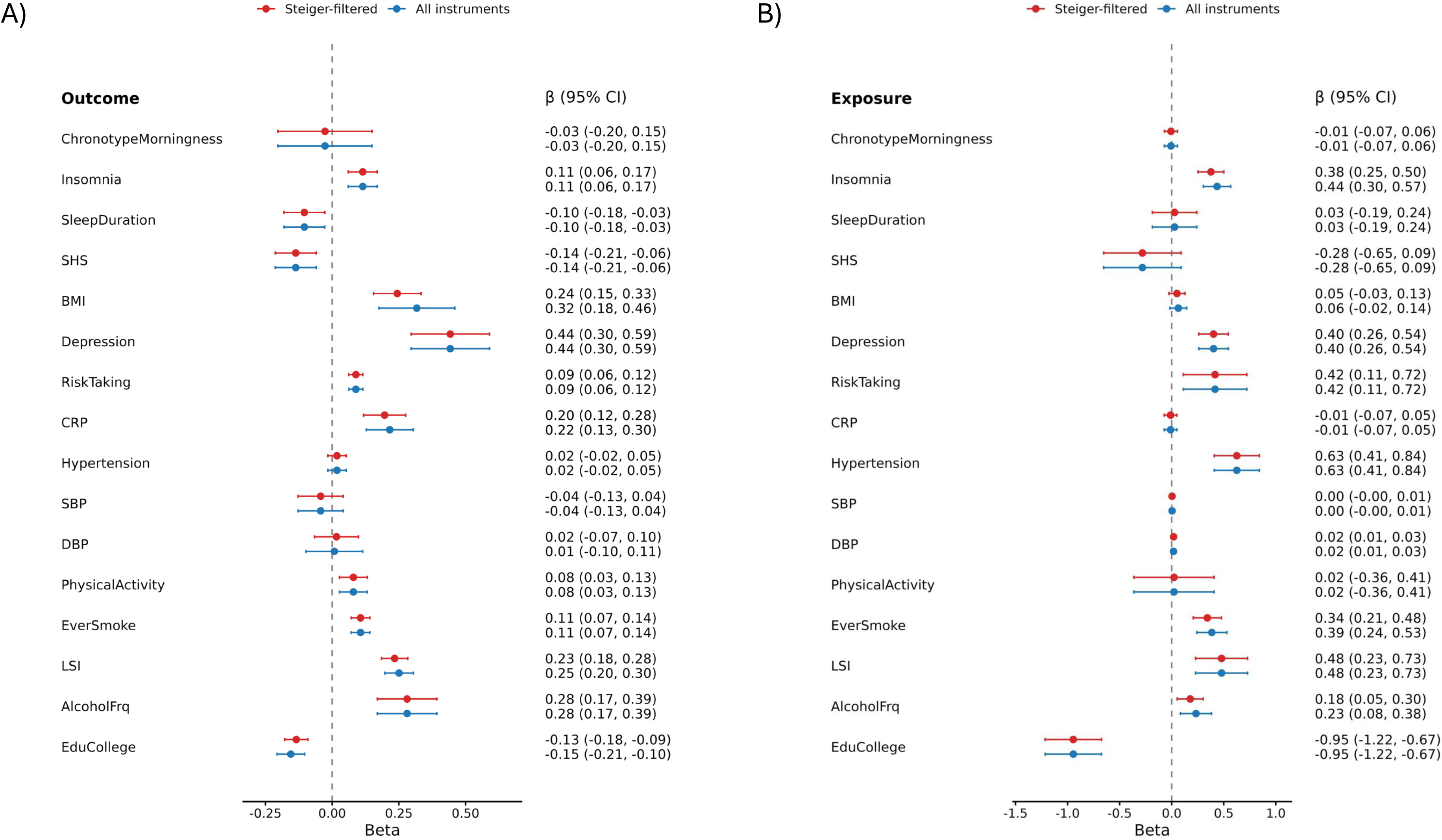
Univariable Mendelian randomization estimates for the associations of ADHD liability with candidate mediators (A) and candidate mediators with migraine (B), comparing all instruments with Steiger-filtered instruments.

### Causal effects of genetic liability to mediators on migraine

The subsequent stage of UVMR evaluated the effect of these factors on migraine risk. Genetically predicted depression was associated with increased migraine risk (β = 0.38,, 95% CI = 0.26, 0.54; P = 1.76 × 10^-7^), as were insomnia (β = 0.44, 95% CI = 0.30, 0.57; P = 7.95 × 10^-11^), hypertension (β = 0.63, 95% CI = 0.41, 0.84; P = 3.19 × 10^-9^), diastolic blood pressure (β = 0.02, 95% CI = 0.01, 0.03; P = 1.45 × 10^-5^), ever smoking (β = 0.39, 95% CI = 0.24, 0.53; P = 1.75 × 10^-7^), lifetime smoking index (β = 0.48, 95% CI = 0.23, 0.73; P = 4.52 × 10^-4^), and alcohol intake frequency (β = 0.28, 95% CI = 0.08, 0.38; P = 0.002). In contrast, genetically predicted college education (β = -0.93, 95% CI = -1.22, -0.67; P = 2.00 × 10^-11^) was associated with lower migraine risk. MR-Steiger directionality filtering did not provide evidence that the identified mediator-migraine pathways were driven by reverse directionality (Figure 2). Sensitivity analyses are presented in Table S7.

### MR based mediation results

Based on the UVMR analysis, seven candidate mediators showed support for both the ADHD-mediator and mediator-migraine pathways. Among these, depression and alcohol consumption frequency showed conditional sufficient instrument strength (conditional F >10, Table S8) and were taken forward for MVMR-based mediation analysis. Depression showed evidence of mediation, accounting for 34.17% of the total effect of genetic liability to ADHD on migraine (95% CI = 6.17% to 61.63%). Alcohol frequency showed a smaller and less precise indirect effect of 0.05 (95% CI: 0.00 to 0.14), corresponding to 14.44% of the total effect (Table 1). In MVMR, the ADHD-migraine relationship persisted after conditioning on depression liability or alcohol frequency, suggesting that these traits may partly explain the association but do not fully account for it (Table S9).

For ever smoking, lifetime smoking index, insomnia, risk taking, and college education, MVMR-based mediation was not feasible or sufficiently instrumented because of weaker conditional instrument strength (Table S8) or because only top variants were publicly available. These mediators were therefore assessed using a two-step UVMR approach as an alternative. Lifetime smoking index and college education showed evidence of indirect effects, accounting for 30.7% (95% CI: 6.96% to 54.43%) and 37.38% (95% CI: 11.64% to 63.13%) of the total effect of genetic liability to ADHD on migraine, respectively. Ever smoking, insomnia, and risk taking accounted for smaller proportions of the total effect (Table 1).

**Table 1.**
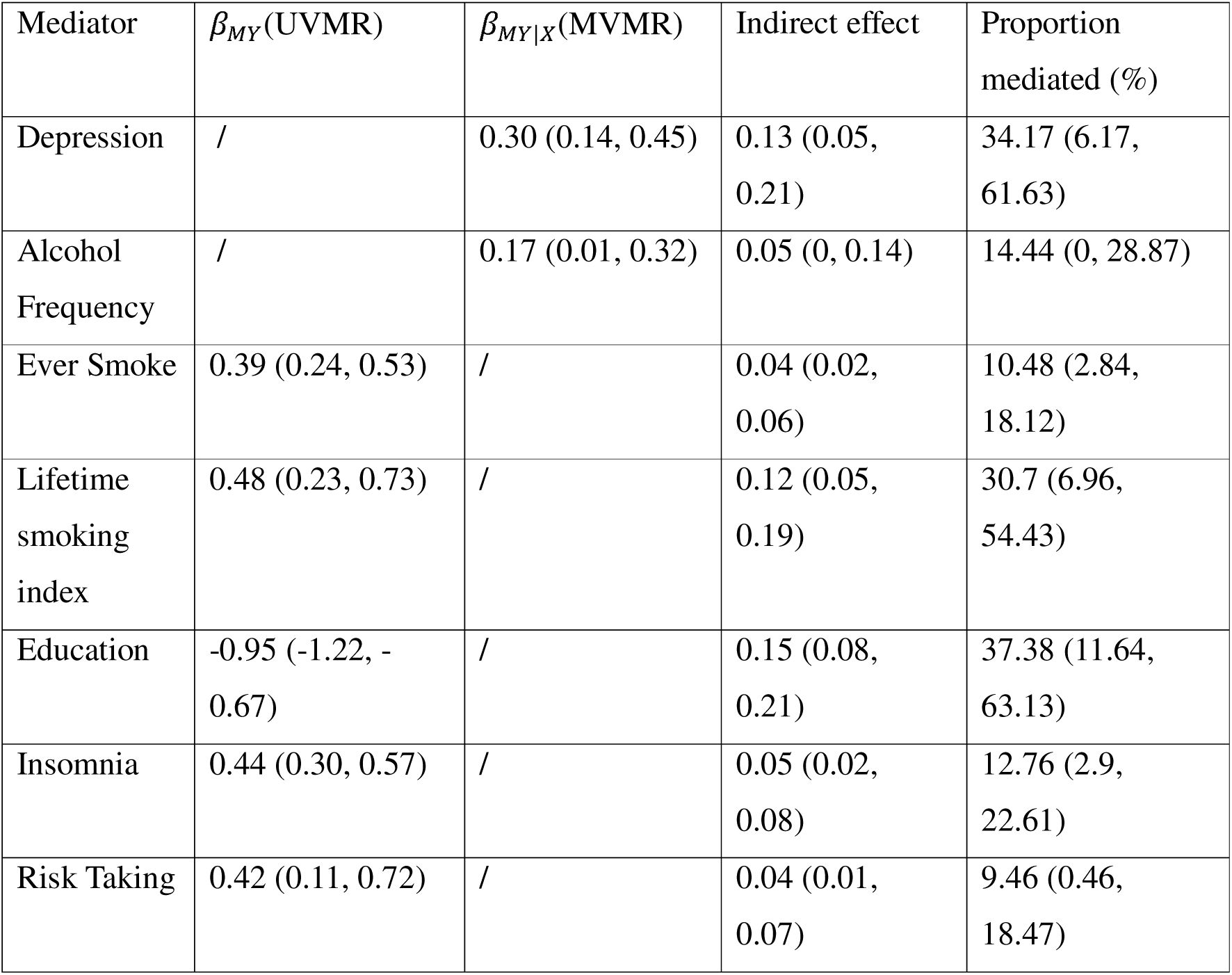
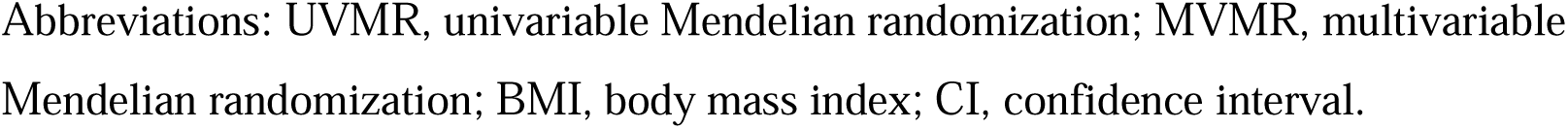
Results of mediation analysis for potential mediation pathway between ADHD and migraine.

| Mediator | $\beta_{MY}(\text{UVMR})$ | $\beta_{MY\vert X}(\text{MVMR})$ | Indirect effect | Proportion mediated (%) |
| --- | --- | --- | --- | --- |
| Depression | / | 0.30 (0.14, 0.45) | 0.13 (0.05, 0.21) | 34.17 (6.17, 61.63) |
| Alcohol Frequency | / | 0.17 (0.01, 0.32) | 0.05 (0, 0.14) | 14.44 (0, 28.87) |
| Ever Smoke | 0.39 (0.24, 0.53) | / | 0.04 (0.02, 0.06) | 10.48 (2.84, 18.12) |
| Lifetime smoking index | 0.48 (0.23, 0.73) | / | 0.12 (0.05, 0.19) | 30.7 (6.96, 54.43) |
| Education | -0.95 (-1.22, -0.67) | / | 0.15 (0.08, 0.21) | 37.38 (11.64, 63.13) |
| Insomnia | 0.44 (0.30, 0.57) | / | 0.05 (0.02, 0.08) | 12.76 (2.9, 22.61) |
| Risk Taking | 0.42 (0.11, 0.72) | / | 0.04 (0.01, 0.07) | 9.46 (0.46, 18.47) |

Abbreviations: UVMR, univariable Mendelian randomization; MVMR, multivariable Mendelian randomization; BMI, body mass index; CI, confidence interval.

*β_MY_* denotes the UVMR estimate for the effect of the mediator on migraine, and *β_MY_*_|*X*_ denotes the MVMR estimate for the effect of the mediator on migraine conditional on ADHD liability. MVMR-based mediation was used where feasible and sufficiently instrumented; otherwise, UVMR-based mediation was used as complementary evidence. Indirect effects were calculated as *β_XM_* × *β_MY_*_|*X*_ for MVMR-based mediation and *β_XM_* × *β_MY_* for UVMR-based mediation. Proportion mediated was calculated as IE/TE, where TE is the total effect of ADHD liability on migraine. A slash indicates that the estimate was not used in the mediation calculation.

### Causal effect of ADHD genetic liability on migraine subtypes

In secondary subtype analyses, the IVW estimates suggested that genetic liability to ADHD was associated with increased risk of both MA (OR = 1.52, 95% CI: 1.18, 1.96, P = 0.001) and MO (OR = 1.65, 95% CI: 1.19, 2.31, P = 0.003). Sensitivity estimates were generally directionally consistent but less precise, with weaker evidence from weighted median, MR-Egger, and mode-based methods (Table S10). There was limited evidence of directional horizontal pleiotropy based on the MR-Egger intercept.

For candidate mediators, subtype analyses showed some overlap with the findings for overall migraine (Table S11-12). Genetic liability to insomnia, depression, hypertension, ever smoking, lifetime smoking index and college education showed evidence of association with both MA and MO. Genetic liability to BMI and alcohol intake frequency showed weak evidence of a positive association with MO, with an estimate close to null for MA. The remaining candidate mediators show little evidence for association with either subtype (Figure 3).

**Figure 3.**
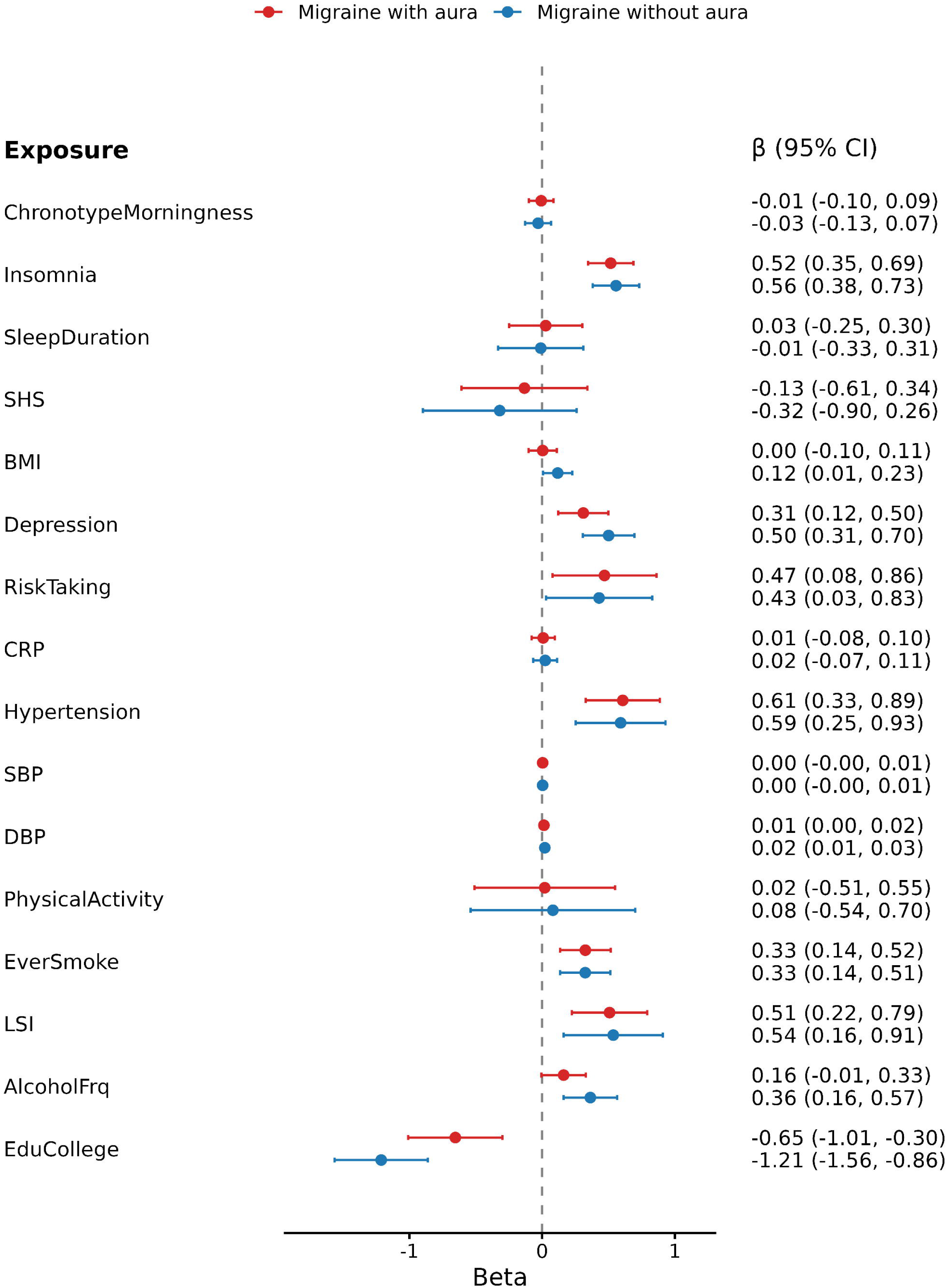
Univariable Mendelian randomization estimates for the associations of candidate mediators with migraine with aura and migraine without aura.

Subtype-specific mediation analyses showed broadly similar patterns for MA and MO. Genetic liability to depression, lifetime smoking index, and education showed evidence of indirect effects for both subtypes, with proportions mediated of 32.14% (95%CI 3.37%, 61.01%), 30.35% (95%CI 4.53%, 56.18%), and 24.10% (95% CI 2.90%, 45.30%) for MA (Table S13), and 37.49% (95% CI 5.5%, 69.47%), 26.72% (95%CI 0.44%, 53.00%), and 37.29% (95%CI 7.65%, 66.94%) for MO (Table S14), respectively. Alcohol intake frequency showed evidence of mediation for MO but not MA, although confidence intervals overlapped, providing limited evidence for a clear subtype difference. BMI was assessed only for MO, as it did not meet the prioritisation criteria for MA because of insufficient evidence for the BMI-MA association.

## Discussion

In this study, we used MR to investigate whether genetic liability to ADHD is associated with migraine risk and to explore the potential mediating pathways. ADHD liability was associated with higher risk of overall migraine, with similar association patterns for both MA and MO. While previous observational and genetic studies have reported overlap between ADHD and migraine (42), our findings extend this evidence by suggesting that psychiatric, behavioural, and socioeconomic pathways may partly contribute to this association.

Among the candidate mediators, the clearest evidence was observed for depression-related liability, lifetime smoking index, and educational attainment. In contrast, BMI, alcohol frequency, and sleep-related traits showed weaker or less consistent evidence in the main mediation analyses. This pattern suggests that the ADHD-migraine association is unlikely to be explained by a single pathway, but may instead reflect a combination of psychiatric, behavioural, lifestyle-related, and educational pathways.

Depression-related liability was one of the clearest potential pathways linking ADHD liability and migraine. This is consistent with previous observational and genetic studies showing close relationships between ADHD, depression, and migraine, including evidence for shared liability between psychiatric traits and migraine (43). Our findings extend this work by suggesting that depression-related liability may partly mediate the effect of ADHD-migraine relationship. Several mechanisms may contribute to the depression-migraine relationship, including altered stress-response systems, pain modulation, and inflammation pathways (44). In MVMR, however, ADHD liability remained associated with migraine after conditioning on depression-related liability, suggesting that depression may explain part, but not all, of the ADHD-migraine relationship.

Smoking-related behaviours also emerges as a plausible behavioural pathway. This is consistent with previous genetic evidence that ADHD liability is closely linked to smoking initiation, persistence, and heaviness (24). Our findings extend this evidence by suggesting that cumulative smoking exposure, indexed by lifetime smoking index, may partly mediate the ADHD-migraine relationship. Smoking may contribute to migraine risk through several ways. Tobacco exposure can impair vascular relaxation and endothelial function, while promoting inflammatory cell activation and oxidative stress (45). These processes may increase neurovascular sensitivity and lower the threshold for migraine attacks (46).

Insomnia may represent a sleep-related pathway linking ADHD liability and migraine. This is consistent with previous evidence that ADHD is commonly associated with difficulties in sleep initiation and sleep maintenance (47). Sleep disturbance may increase migraine susceptibility through altered pain processing and stress-response activation, and increased susceptibility to cortical spreading depression, a neurophysiological process implicated in migraine (48). Our findings extend this evidence by suggesting that insomnia may partly mediate the ADHD-migraine relationship, although this pathway was supported by UVMR-based mediation and could not be assessed using MVMR.

Educational attainment was also supported in the mediation analyses, but its interpretation is less straightforward. This finding is consistent with previous evidence linking ADHD liability to educational outcomes and with studies showing that the education is socially and genetically correlated with a wide range of health-related traits (49, 50). Educational attainment is unlikely to represent a single biological mechanism; rather, it may capture broader cognitive, socioeconomic, developmental, and family-level processes. The observed mediation effect may therefore reflect these broader education-related pathways rather than a specific causal mechanism operating through educational attainment itself.

Subtype-specific analyses suggested broadly similar patterns for MA and MO. The total effect estimates for ADHD liability were similar across subtypes, and the mediation patterns were largely overlapping. This suggests that ADHD liability and the examined mediating pathways may contribute to migraine susceptibility across subtypes. BMI was taken forward only for MO because there was evidence for BMI-MO association but limited evidence for a BMI-MA association. Given that previous observational evidence has linked obesity to both MA and MO, with similar association reported across aura-defined subtypes (51), this BMI result should be viewed as a tentative metabolic pathway requiring further investigation rather than clear evidence of subtype specificity.

This study has several strengths. First, we used a genetically informed design to examine the relationship between ADHD liability and migraine, reducing the potential influence of reverse causation and some forms of environmental confounding that may affect observational studies. Second, we applied a mediation framework to evaluate several behavioural, psychiatric, sleep-related, educational, and cardiometabolic traits as potential pathways, allowing us to identify which downstream traits may contribute to the ADHD-migraine association. Third, where instrument strength allowed, MVMR was used to estimate mediator effects conditional on ADHD liability, helping to distinguish potential mediators from correlated genetic liability. Finally, by examining MA and MO separately, we extended previous evidence based on overall migraine and explored whether ADHD-related pathways differed across clinically relevant migraine subtypes.

Several limitations should also be considered. First, causal inference from MR estimates relies on the core instrumental variable assumptions, and horizontal pleiotropy cannot be fully excluded, particularly for complex psychiatric and behavioural traits. Second, several candidate mediators were strongly genetically correlated with ADHD liability, which limited the use of MVMR because of weak conditional instrument strength. Third, mediation MR may oversimplify complex developmental and behavioural processes. Traits such as education and smoking-related behaviour may partly index broader developmental, or externalising liabilities that are correlated with both ADHD liability and migraine. Because these candidate mediators are genetically correlated with ADHD liability, MVMR analyses had limited ability to estimate whether their effects on migraine were independent of ADHD liability. Finally, the analyses were based largely on individuals of European ancestry, which may limit generalisability to other populations.

In conclusion, our findings provide evidence that ADHD liability is associated with increased risk of migraine, with similar evidence observed for both MA and MO. The mediation results suggest that this association may involve depression-related liability, smoking-related behaviour, and broader education-related processes. Some subtype-specific patterns were observed, although these were imprecise and should be interpreted with caution. Together, these findings support a multifactorial model of ADHD-migraine comorbidity and suggest that future work should consider both downstream behavioural and psychiatric pathways and heterogeneity between migraine subtypes.

## Ethical Approval

No new ethical approval was required for this study because all analyses were conducted using publicly available, anonymised GWAS summary statistics. Ethical approval and informed consent had been obtained by the investigators of the original studies from which the summary statistics were derived.

## Data Availability

All data used in this study are publicly available from the sources described in the Methods and Supplementary Materials.

